## Extended Data Figures for "Single-cell genomics improves the discovery of risk variants and genes of Atrial Fibrillation"

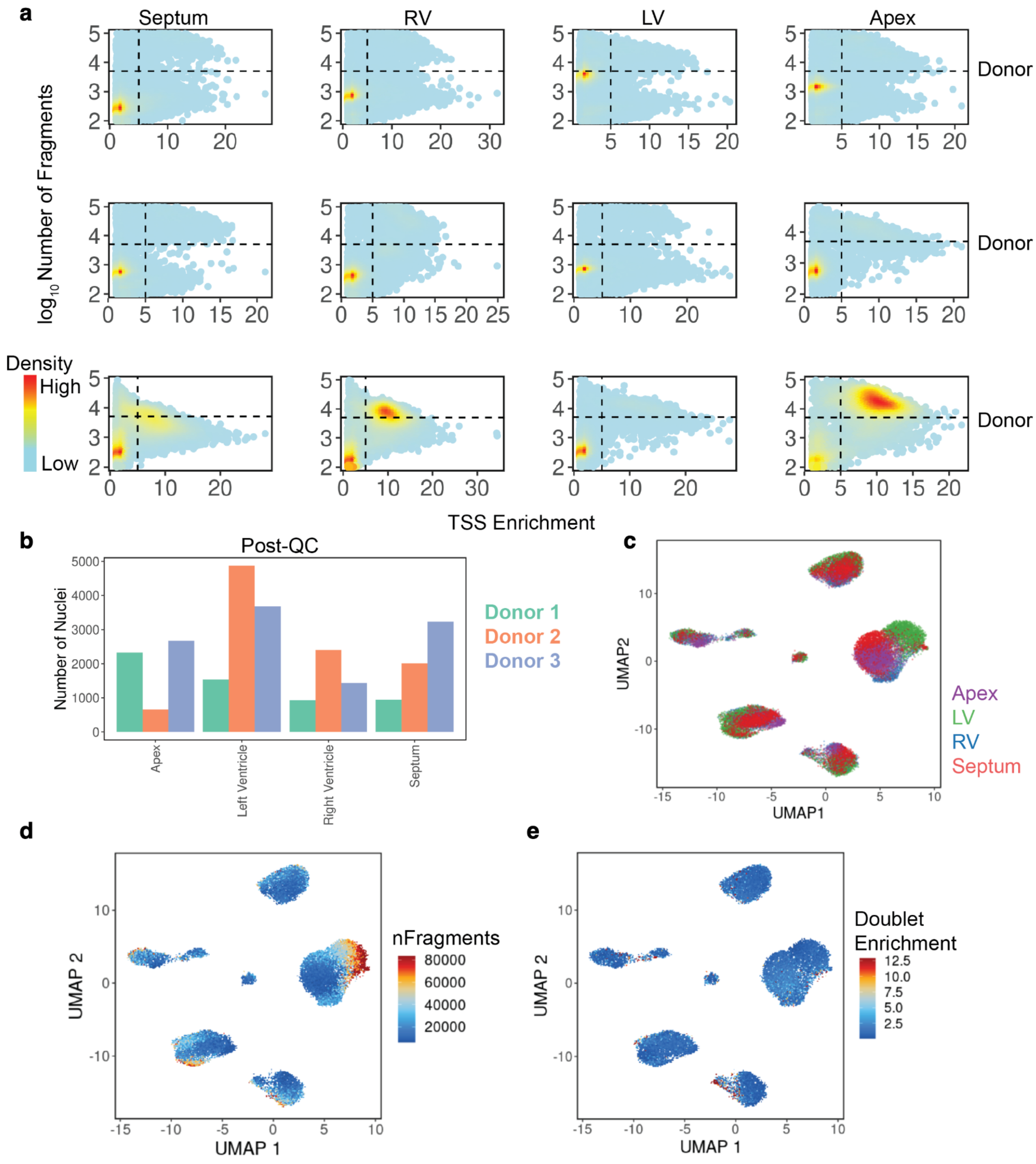


**Extended Data Fig. 1: QC metrics for scATAC-seq samples across samples, donors, and clusters. a**, Number of reads vs. TSS enrichment for all included libraries. Cells with more than 5,000 unique fragments and a TSS enrichment score above 5 were included. **b**, Number of cells passing QC per region and donor. **c**, Anatomic location, **d**, Number of fragments, **e**, Doublet enrichment scores overlaid on two-dimensional representation of cells using UMAP.

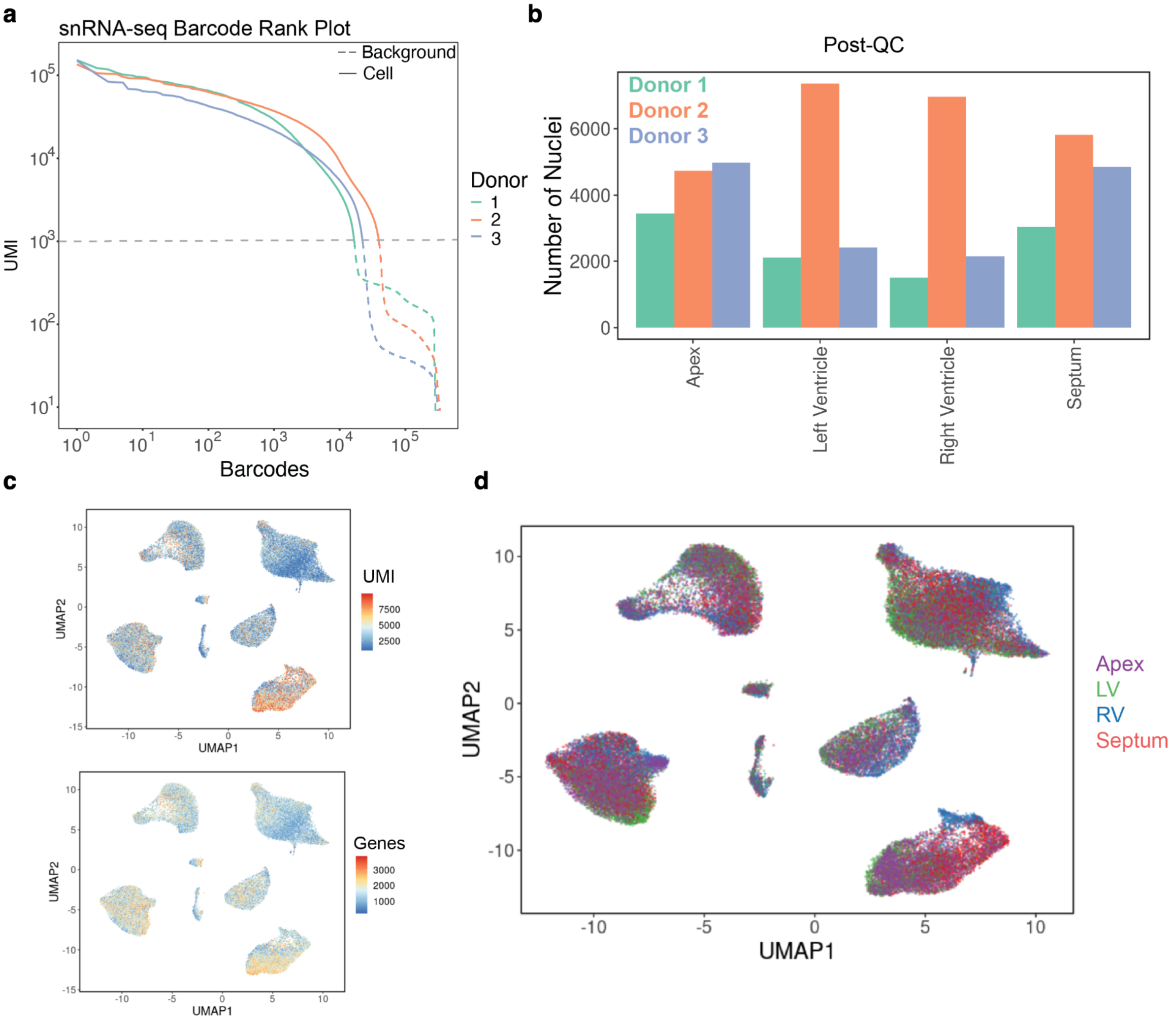


**Extended Data Fig. 2: QC metrics for snRNA-seq samples across samples, donors, and clusters. a**, UMIs vs barcodes ranked by UMI count. Shown are combined samples for each donor. Cells with more than 1,000 UMIs were included. **b**, Number of nuclei passing QC per region and donor. **c**, Number of UMIs (top) and detected genes and **d**, Anatomic location overlaid on two-dimensional representation of nuclei using UMAP.

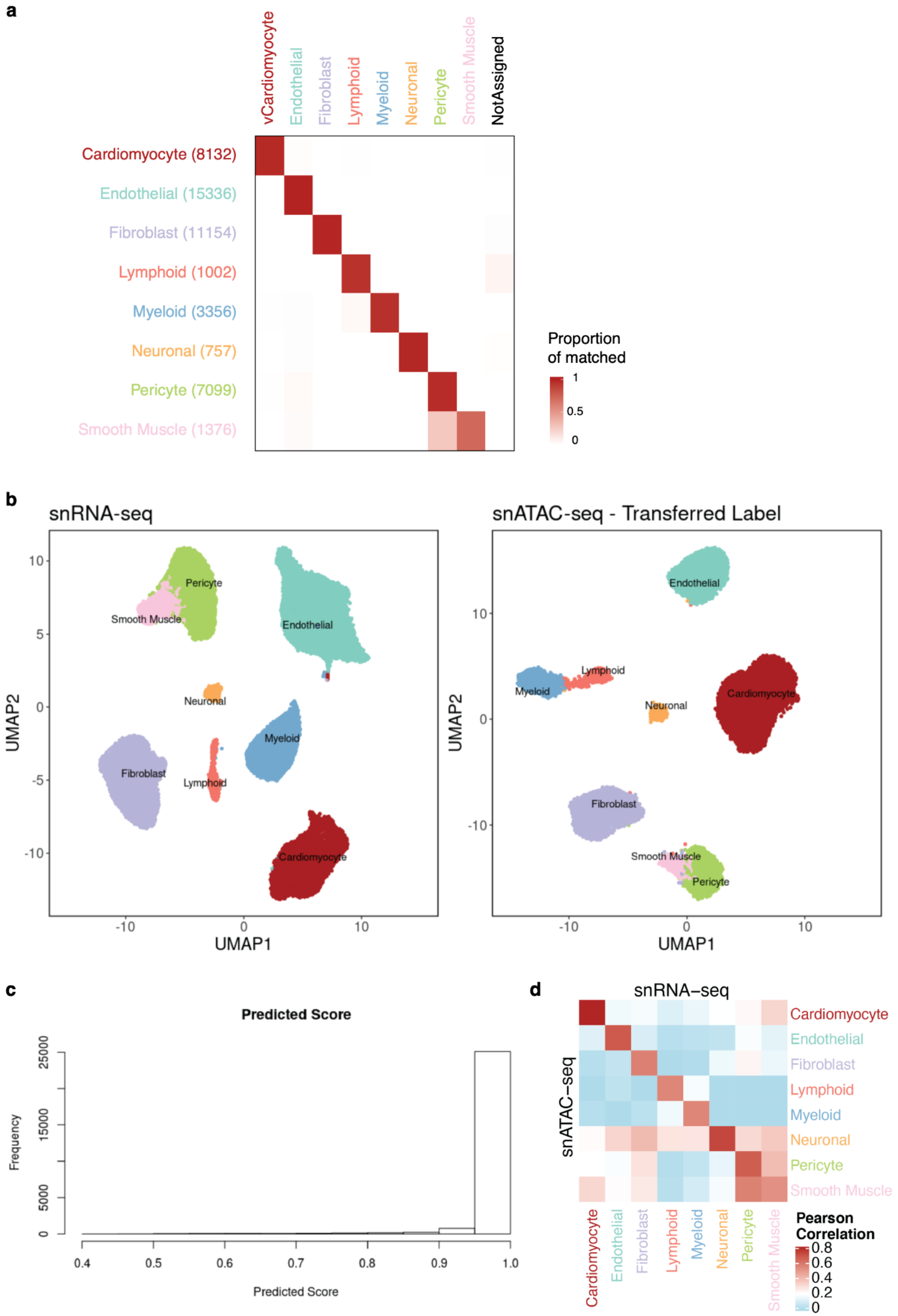
a
vCardiomyocyte
Endothelial
Fibroblast
Lymphoid
Myeloid
Neuronal
Pericyte
Smooth Muscle
NotAssigned
Cardiomyocyte (8132)
Endothelial (15336)
Fibroblast (11154)
Lymphoid (1002)
Myeloid (3356)
Neuronal (757)
Pericyte (7099)
Smooth Muscle (1376)
Proportion of matched
1
0.5
0
b
snRNA-seq
snATAC-seq - Transferred Label
UMAP2
UMAP1
Pericyte
Smooth Muscle
Endothelial
Neuronal
Myeloid
Fibroblast
Lymphoid
Cardiomyocyte
c
Predicted Score
Frequency
d
snRNA−seq
snATAC−seq
Pearson Correlation
0.8
0.6
0.4
0.2
0

**Extended Data Fig. 3: Label transfer of cell-type labels from snRNA-seq to scATAC-seq using Seurat anchor transfer. a,** Comparison of cell-type annotation in our study to labels transferred from Heart Cell Atlas, Litvinukova *et al*.[1]. Rows are cluster labels in our study with the number of cells indicated for each cell type in brackets. Columns are labels from Litvinukova *et al*. Shown are the proportions of cells per cluster (row) that are annotated with a given label in Litvinukova et al. Of the 49539 cells in our study, 48359 (97.6%) were annotated the same labels with Litviňuková's. 1180 (2.4%) had discordant cell type levels and 349 of them were not matched and fell into the "NotAssigned" category. **b**, Heart cells profiled by snRNA-seq (left) and scATAC-seq (right) embedded in two dimensions (UMAP). Cluster labels determined based on transcriptional signatures of snRNA-seq data (left) were transferred to scATAC-seq dataset (right). **c**, Distribution of cluster label prediction scores for each cell in the scATAC-seq dataset. **d**, Pairwise correlations (Pearson's r) between gene scores (scATAC-seq) and transcript levels (snRNA-seq) for differentially expressed genes between cell types using aggregated data from all cells within a cluster (pseudo-bulk).

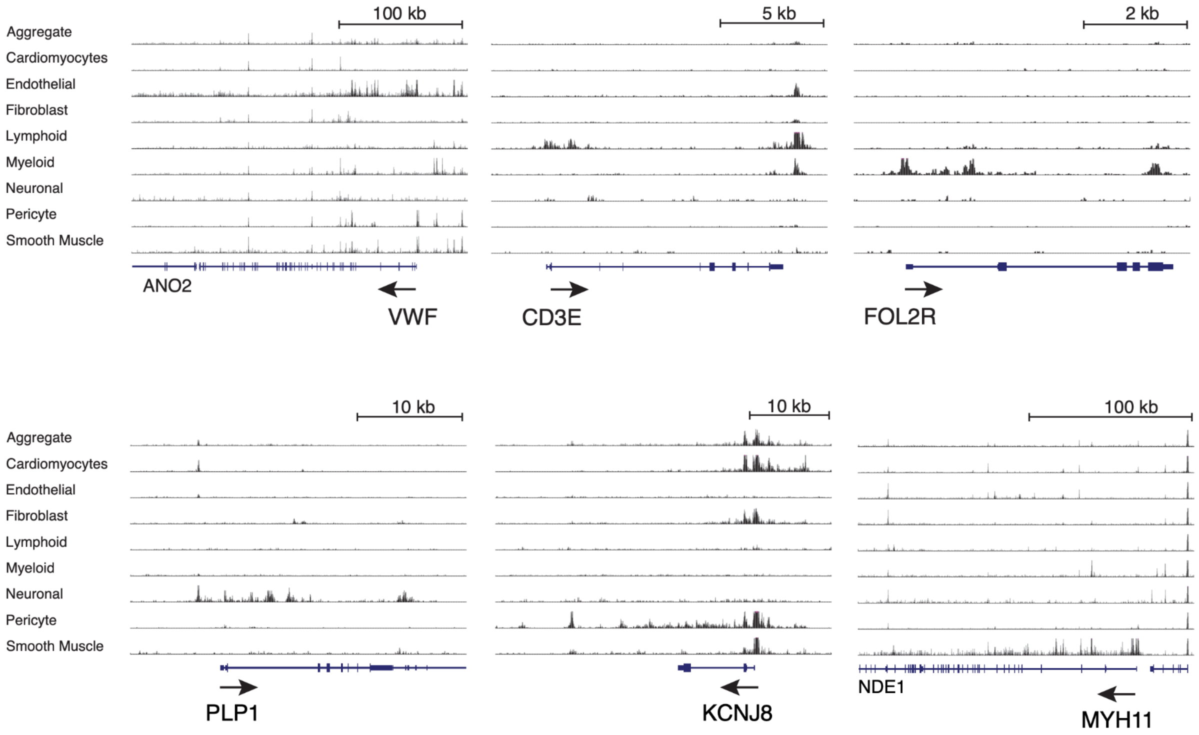


**Extended Data Fig. 4: Genome browser track plots of chromatin accessibility at marker genes across cell types, similar to Figure 2b.**

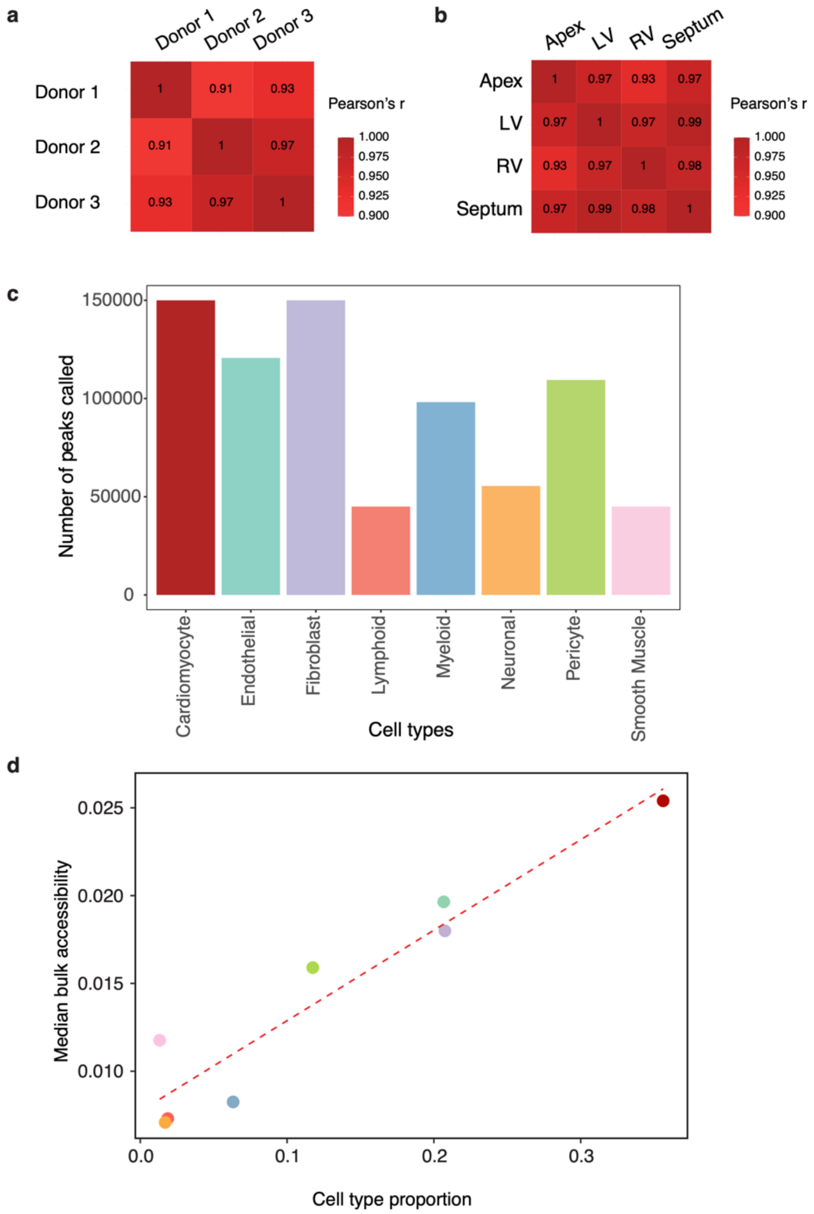


**Extended Data Fig. 5: Comparison of OCRs between individuals and across cell types. a**, Pairwise correlations (Pearson's r) of chromatin accessibility between individual donors. The comparisons were made between pseudo-bulk samples aggregating all cells from each donor. **b**, Pairwise correlations (Pearson's r) of chromatin accessibility between sampling locations. The comparisons were made between pseudo-bulk samples aggregating all cells from each location (including all three donors). **c**, Number of detected peaks in each cluster/cell type using MACS2. **d,** Correlation between the proportion of a cell type in the heart (X-axis) and the median accessibility, at the bulk level, of all peaks found in that cell type (Y-axis). Each dot represents one cell type, using the color scheme of Panel c). To define the median accessibility signal of a cell type, we first obtained the accessibility signal of any peak from the pseudo-bulk data, pooling all cells. We then took the median of the accessibility of all peaks called in any given cell type.

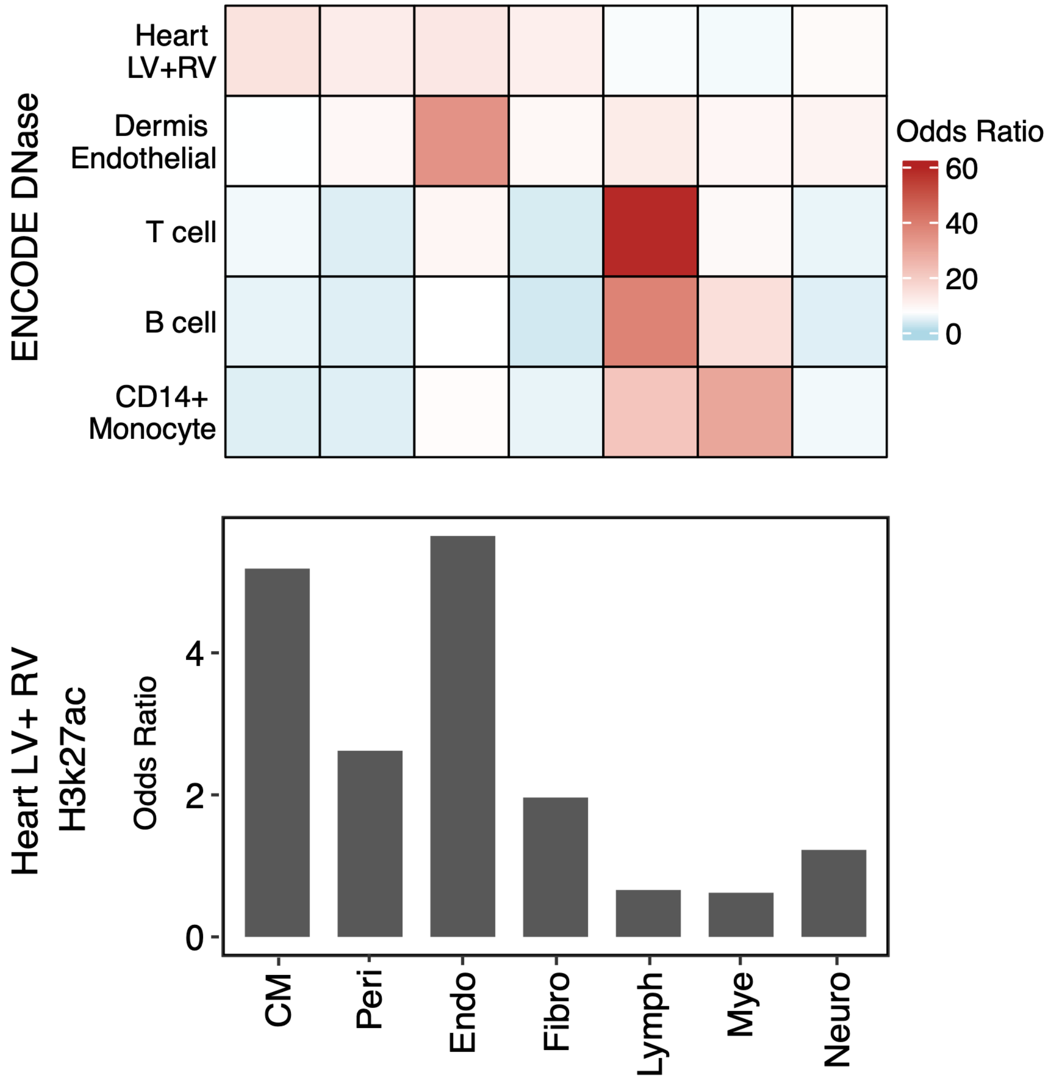


**Extended Data Fig. 6: Comparison of open chromatin regions between the current study and ENCODE DNase hypersensitivity sites (DHS) from the indicated tissues (a) and ENCODE H3K27ac peaks in the heart (b).**

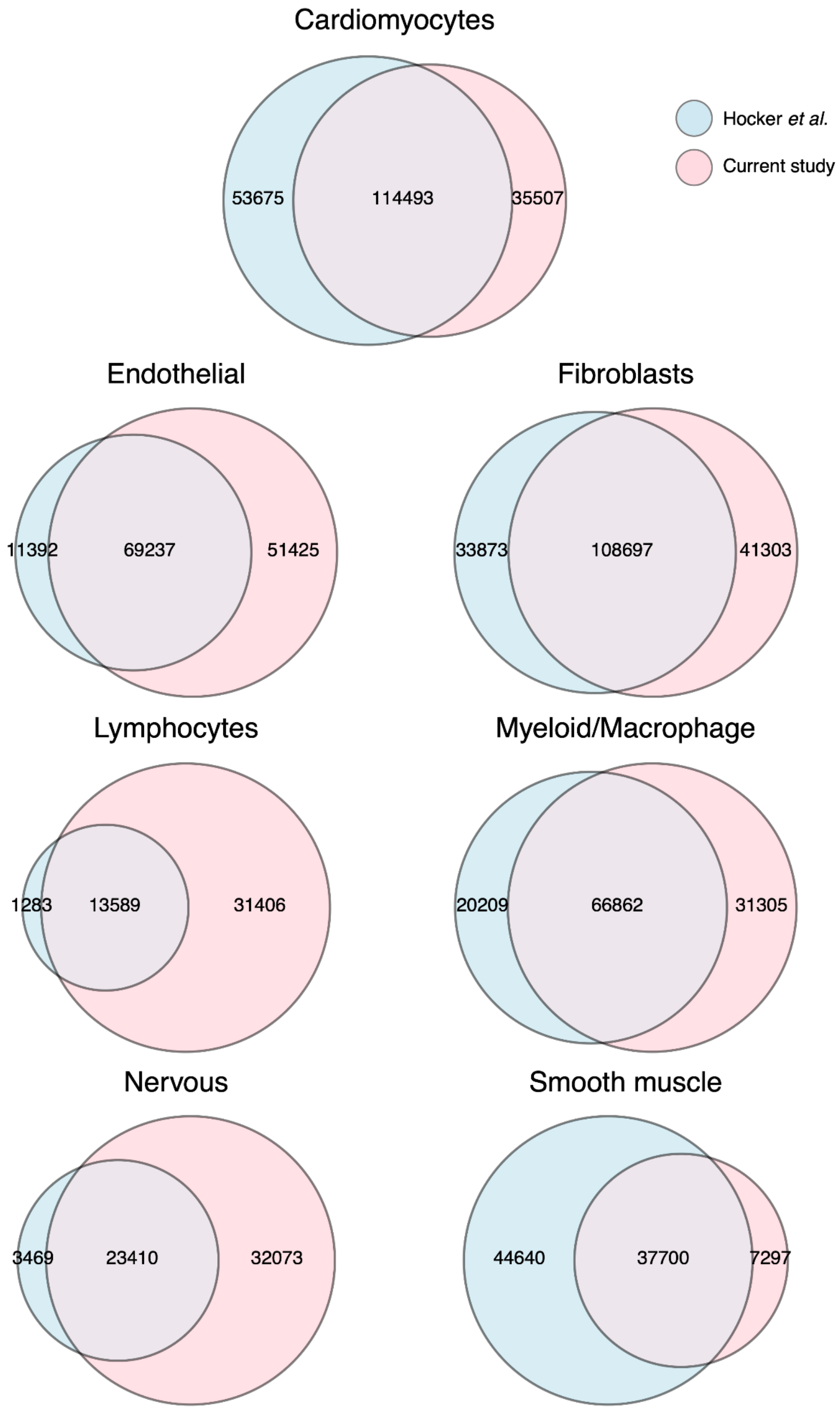


**Extended Data Fig. 7: Comparison of Peaks identified by Hocker *et al.* and this study.** The numbers of peaks in the overlapped areas of the Venn Diagrams are from this study. The numbers of overlapped peaks from Hocker *et al.* are similar.

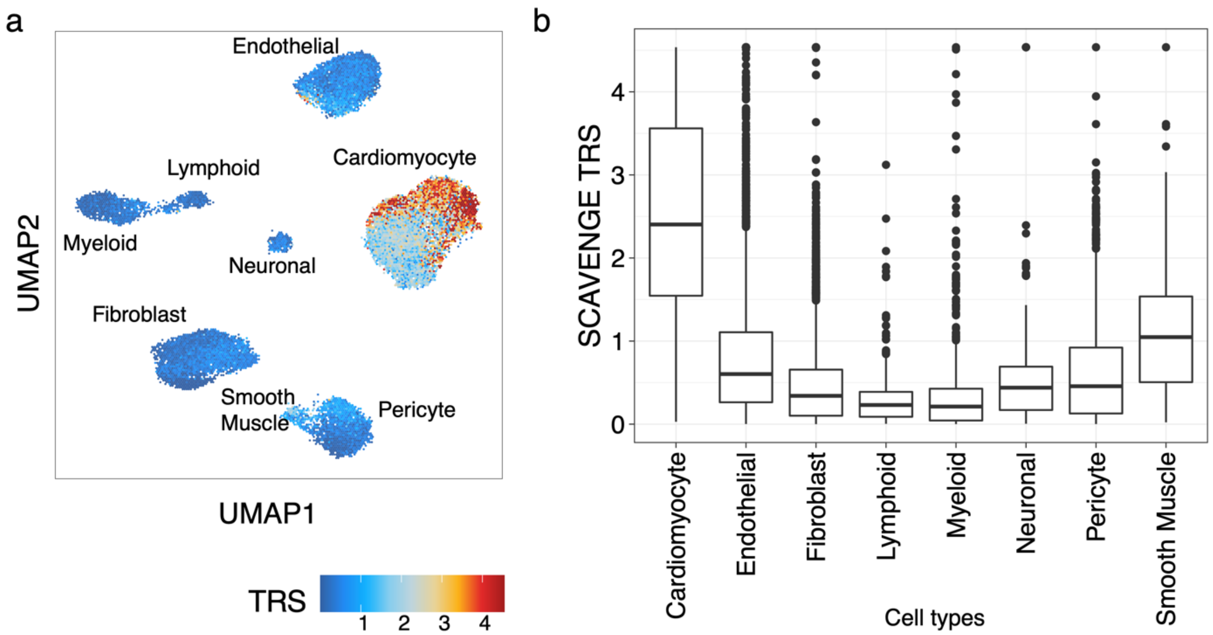


**Extended Data Fig. 8: TRS scores from SCAVENGE are highest in Cardiomyocytes.** (a) TRS for each cell plotted on UMAP embedding. (b) Distribution of TRS for each of our cell type clusters.

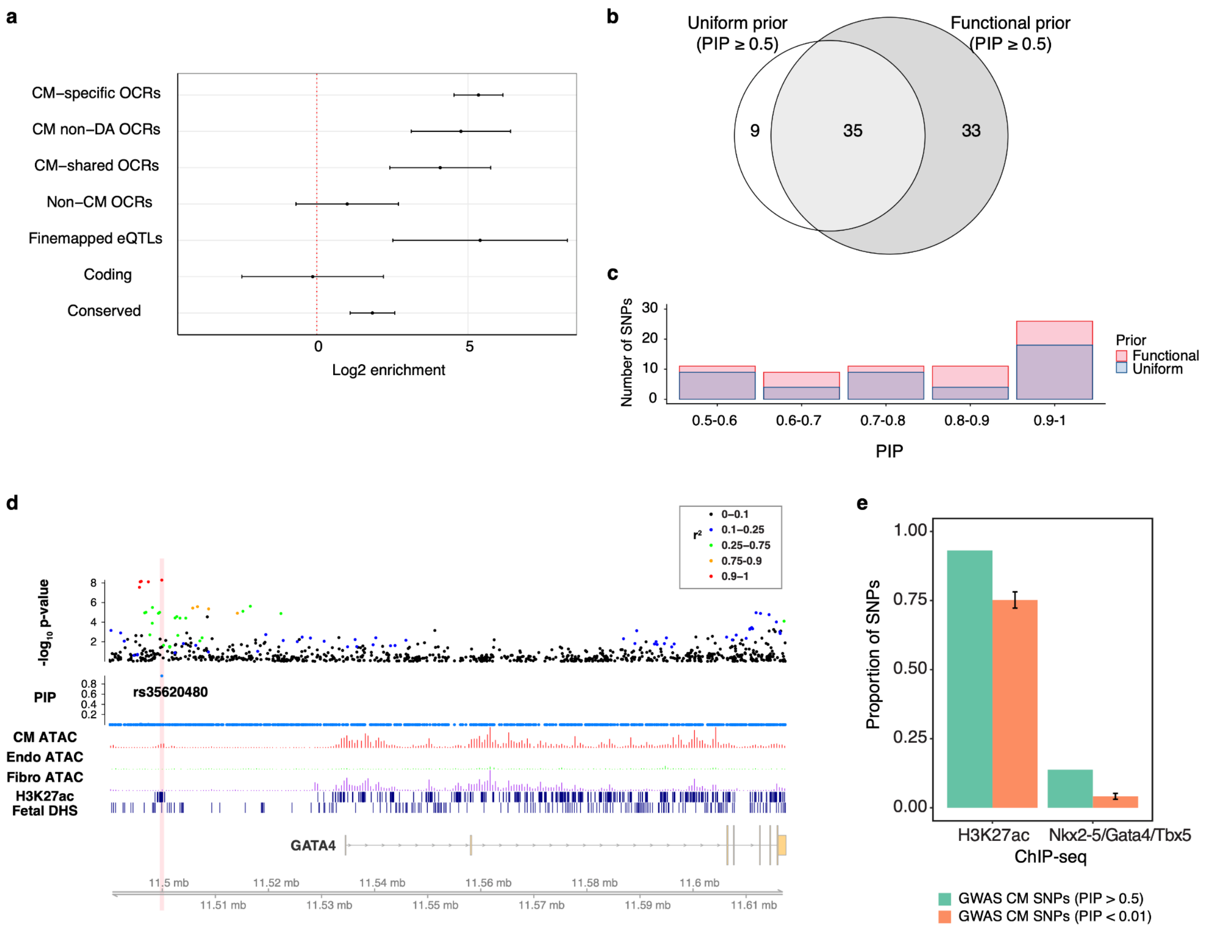


**Extended Data Fig. 9: Enrichment analysis and function of putative AF risk variants. a**, Log2 fold enrichment of AF risk variants in five functional categories, based on TORUS. The enrichment values were obtained in a joint model where all five annotations are included. The resulting variant probabilities from TORUS were used as priors for fine-mapping with SuSiE. **b,** SNPs with PIP >= 0.5 identified using functional prior vs. uniform prior **c,** Comparing PIP distributions between functional prior and uniform prior (only showing SNPs with PIP >= 0.5). **d**, Example of fine mapping with functional prior around GATA4 locus. **e**, Proportions of high and low confidence SNPs (based on fine-mapping) in H3K27ac ChIP-seq region in the human heart (ENCODE) and in Nkx2-5/Gata4/Tbx5 ChIP-seq regions identified in the mouse heart and lifted over to the human genome.

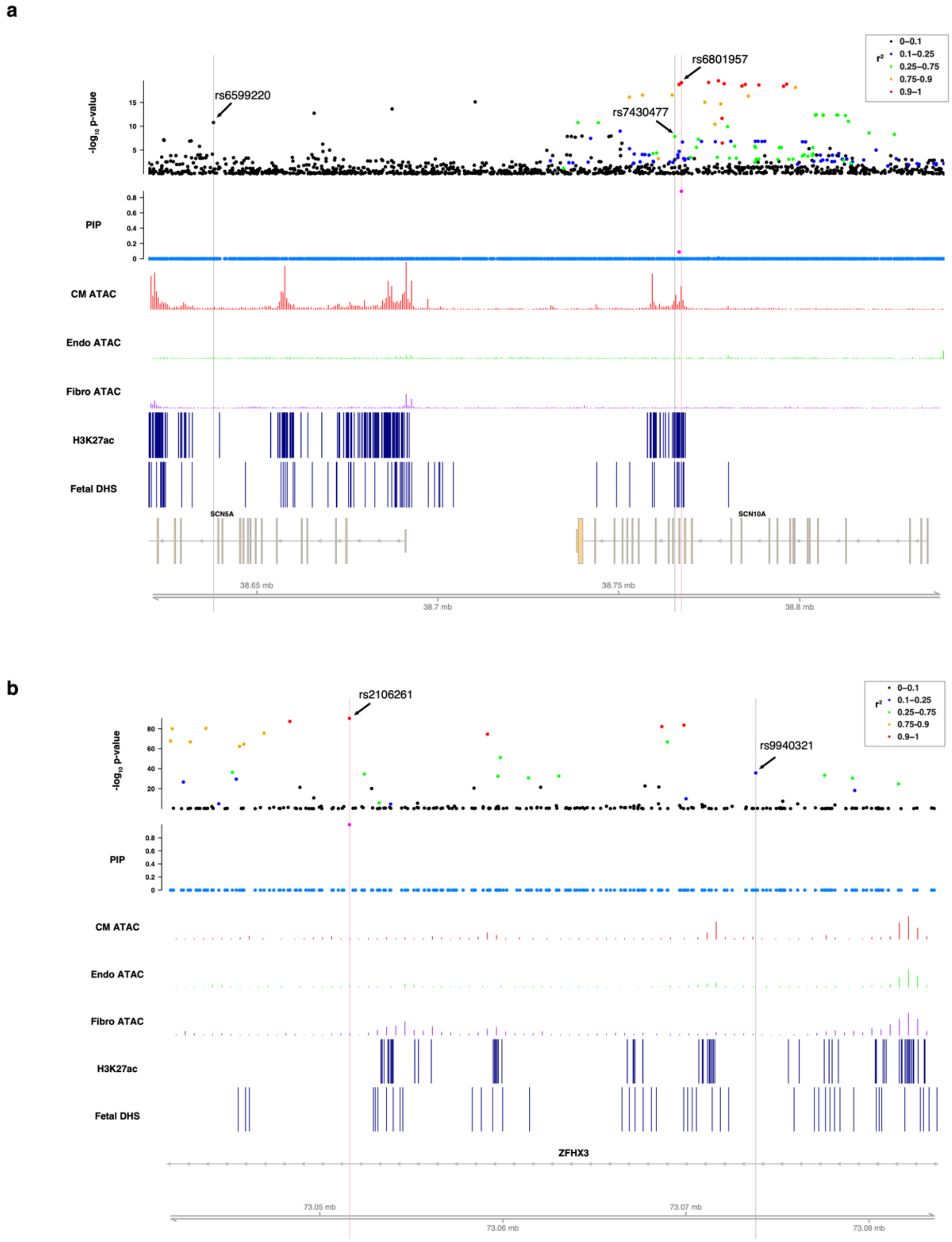


**Extended Data Fig. 10: Examples of fine-mapping results for allele-specific variants from STARR-seq.** For each locus, data tracks shown (from top to bottom) are: association with AF phenotype from GWAS; PIP from fine mapping taking into account OCRs and other functional categories; pseudo-bulk chromatin accessibility profiles for CMs (red), endothelial cells (green), and fibroblasts (purple); H3K27ac ChIP-seq and fetal DHS peak calls. Allele-specific variants from STARR-seq (van Ouwerkerk et al., Circ. Res. 2020) were highlighted in gray, and fine-mapped variants from the current study were highlighted in pink.

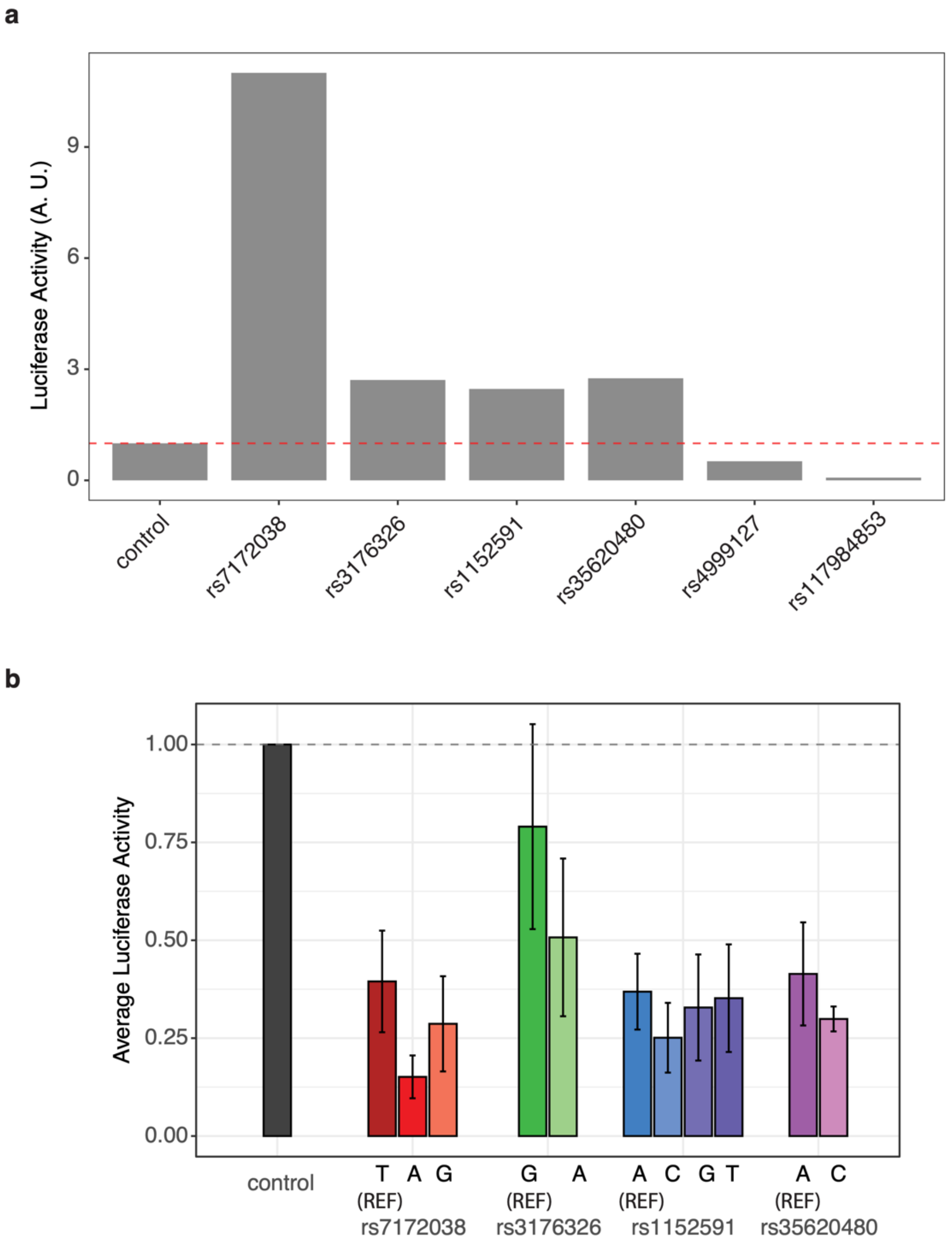


**Extended Data Fig. 11: Assessing reporter gene activation for selected regions and variants. a,** Result of initial screening of 6 candidate regions for reporter gene activity. Experiment was used to filter out regions or constructs that showed no activity before moving to measurements of allelic effects. Only a single replicate was obtained at this stage. **b,** Reporter activities in 3T3 fibroblasts of regions containing selected SNPs, with reference and alternative alleles (same regions as in Fig. 4j).

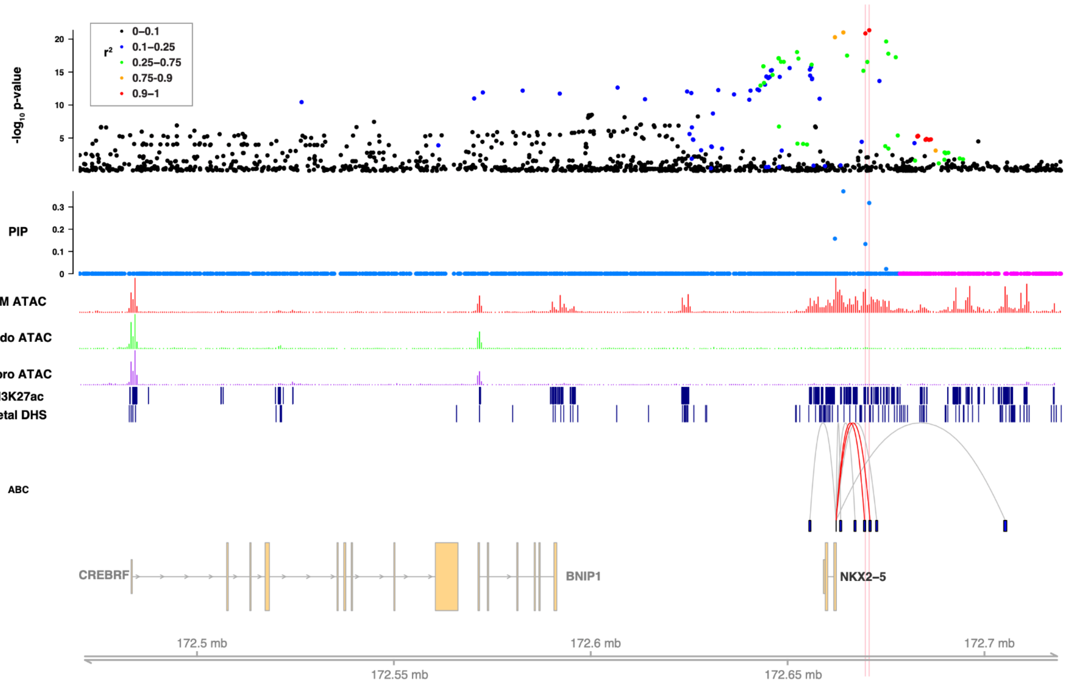


**Extended Data Fig. 12: An example locus containing *NKX2-5* as the candidate gene (PIP = 0.99).** Data tracks shown (from top to bottom) are: association with AF phenotype from GWAS; PIP from fine mapping taking into account OCRs and other functional categories; pseudo-bulk chromatin accessibility profiles for CMs (red), endothelial cells (green), and fibroblasts (purple); H3K27ac ChIP-seq and fetal DHS peak calls; interaction data from ABC scores from heart ventricle. Interactions anchored on the highlighted SNPs are highlighted in red.

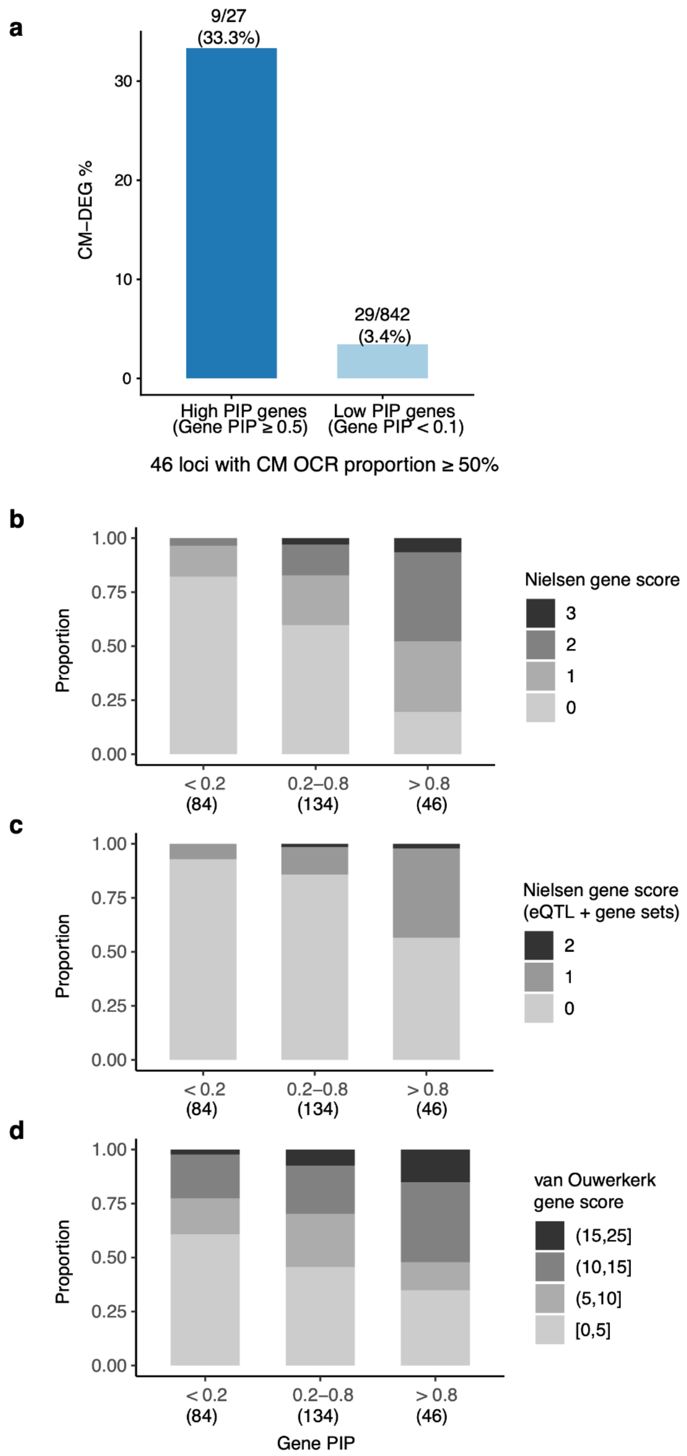


**Extended Data Fig. 13: a, Enrichment of CM DEGs in high PIP genes vs. low PIP genes in loci with high CM OCR proportion. b,c,d, Support of high confidence genes (gene PIP >= 0.8) from functional data collected in earlier AF studies. b,** The proportions of genes at different gene PIP bins with a functional score (0-3) assigned by Nielsen *et al.*[2] The score of a gene was calculated from four sources of information about the GWAS lead SNP and the gene: distance of SNP to gene, exonic variant, eQTL and gene sets. A gene thus has a score of 0-4, with each category contributing a score of 1. **c**, as in (**b**) but the scores were calculated based only on eQTL and gene set annotations. This is to reduce the bias that was introduced in computing gene PIPs, i.e. genes with exonic variants and closest to GWAS SNPs tend to have higher PIPs. **d**, as (**b**) but using gene scores developed in van Ouwerkerk *et al.*[3] Genes that are in the same Topologically Associating Domains (TADs) of the GWAS SNPs, eQTL targets, and expressed in heart tissues across multiple datasets have high scores.

a

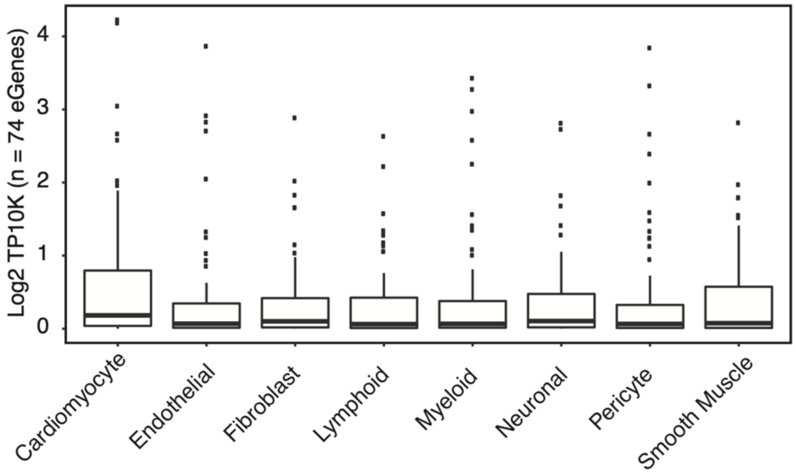


b

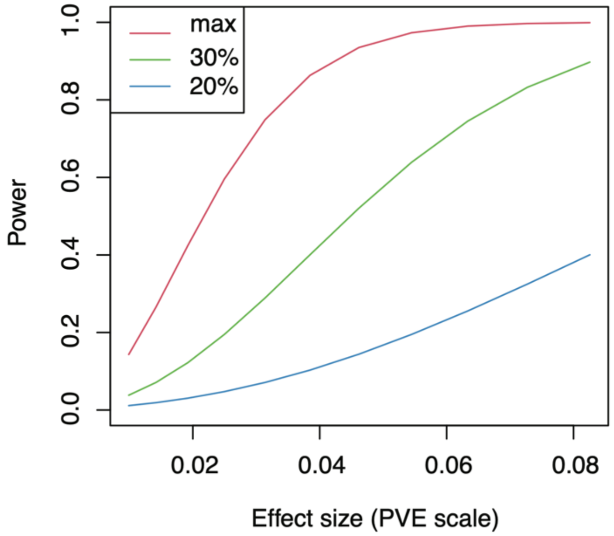


**Extended Data Fig. 14: Tissue sharing analysis of eQTLs. a,** Expression levels of 74 eGenes associated with CM-specific OCRs in heart cell types, from scRNA-seq. See Supplementary Notes. **b**, Power of detecting cell-type-specific eQTLs in bulk samples, as function of effect size. Simulated are 3 scenarios reflecting different proportions with which a cell type occurs in the bulk tissue (1, 0.5, 0.3). The effect size of an eQTL is defined at the PVE scale, i.e. the proportion of variance of gene expression explained by the genetic variation at that eQTL.
