## Supplemental Notes for "Single-cell genomics improves the discovery of risk variants and genes of Atrial Fibrillation"

### Power analysis of bulk eQTL detection

Most eQTL studies were performed using bulk samples, which are mix of multiple cell types. Intuitively, eQTLs with effects in common cell types are easier to detect, so are those with effects shared across multiple cell types. We will quantitatively analyze how the power of bulk eQTL study depends on cell type compositions and effect sizes across cell types. A primary goal is to understand how the power of detecting eQTLs acting on single cell types is reduced, compared to the eQTLs with shared effects across cell types.

Let's consider one SNP-gene pair a time. Let $y_i$ be the total expression, across $K$ cell types, of the gene in sample $i$, and $G_i$ be the genotype of sample $i$, $1 \leq i \leq n$. Also let $p_{ik}$ be the proportion of cell type $k, 1 \leq k \leq K$ in sample $i$, with $\sum_k p_{ik} = 1$. The total expression $y_i$ is the sum of expression across $K$ cell types, weighted by the cell type proportions. Let $y_{ik}$ be expression of the gene in cell type $k$ in the sample $i$, we then have:

$$y_i = \sum_k p_{ik} y_{ik}. \tag{1}$$

To relate gene expression with the genotype, we denote $\beta_k$, the effect size of the SNP on gene expression in cell type $k$, which may vary across cell types. Thus for a cell type $k$, we have

$$y_{ik} = \mu_k + G_i \beta_k + \epsilon_{ik} \qquad \epsilon_{ik} \sim N(0, \sigma_k^2), \tag{2}$$

where $\mu_k$ is the average expression in cell type $k$, and $\sigma_k$ its standard deviation. Summing over all cell types, we have the following model of the bulk gene expression in sample $i$:

$$y_i = \sum_k p_{ik} \mu_k + G_i \sum_k p_{ik} \beta_k + \sum_k p_{ik} \epsilon_{ik} \qquad \epsilon_{ik} \sim N(0, \sigma_k^2). \tag{3}$$

To gain some intuitions of how the power may depend on various parameters, we make the simplifying assumption that cell type compositions are similar across samples, i.e. $p_{ik} \approx p_k$, for all $i$. We now have:

$$y_i = \mu + G_i \sum_k p_k \beta_k + \sum_k p_k \epsilon_{ik} \qquad \epsilon_{ik} \sim N(0, \sigma_k^2), \tag{4}$$

where $\mu = \sum_k p_k \mu_k$ is the average gene expression. Comparing this with regression equation in standard eQTL mapping, we see that the effective bulk eQTL effect is simply $\tilde{\beta} = \sum_k p_k \beta_k$. Now we can see that if an eQTL is specific to a single cell type, say, $k$, then the "effective" eQTL effect size becomes $p_k \beta_k$. Clearly, the reduction of effect size is large for rare cell types.

While the change of effect size is straightforward, the problem is complicated by the fact that the power of mapping eQTL depends not only on the effect size, but also the total variance of gene expression. In fact, the power is determined by the sample size and the percent of variance of gene expression explained (PVE) by the genetic variant. The total variance, and hence PVE, according to the model above, depend on the variance of the error term, $\sigma_k$'s, and also on how the errors across cell types may be correlated. Below, we consider several simplified cases, with the goal of comparing power of detecting cell-type shared vs. cell-type specific eQTLs.

We first consider a special case where the eQTL effects are shared across all cell types, i.e. $\beta_k = \beta$ for all cell type $k$. Furthermore, the gene expression have the same variance, and are fully correlated across all cell types. Under this "full sharing" scenario, an eQTL would have exactly the same effect across all cell types. Thus the PVE of the variant on any cell type would be the same as the PVE in the bulk sample. As a result, the power of detecting this shared eQTL would be the same as the power when we have expression data from pure cell types.

Next we consider the case where eQTL is cell type-specific. Suppose $\beta_c \neq 0$ for some cell type $c$, and 0 for all other ones. The effect size in the bulk data is then $\tilde{\beta} = p_c \beta_c$. As explained above, the actual power would depend on expression in other cell types. In the special case where the gene is expressed only in cell type $c$, then the variance of gene expression in other cell types would be 0. The residual variance of gene expression (after considering eQTL effect) is then:

$$\mathrm{Var}\left(\sum_k p_k \epsilon_{ik}\right) = p_c^2 \sigma_c^2. \tag{5}$$

Assuming genotype $G_i$ is standardized i.e. its variance is equal to 1, the PVE of the eQTL on the bulk gene expression is then:

$$\mathrm{PVE} = \frac{p_c^2 \beta_c^2}{p_c^2 \beta_c^2 + p_c^2 \sigma_c^2} = \frac{\beta_c^2}{\beta_c^2 + \sigma_c^2}, \tag{6}$$

which is exactly the PVE of the eQTL in the cell type $c$ alone. This analysis thus suggests that there is no power loss of detecting cell-type specific eQTLs when gene expression is limited to that cell type.

In practice, however, even if an eQTL is specific to a cell type, the associated gene may be expressed much more broadly. Using our scRNA-seq data, we assessed the expression pattern of the eGenes detected in GTEx heart whose eQTLs are fine-mapped to Cardiomyocyte (CM)-specific OCRs. While the eQTL effects are likely CM-specific, the expression of genes are generally not restricted to CMs. In fact, the distributions of expression levels of these genes overlap considerably across cell types (Extended Data Fig. 14a). For example, the median gene expression in CMs corresponds to top 34% expression of endothelial genes, and 38% of fibroblast. Motivated by this observation, and for mathematical simplicity, we assume equal expression variance across cell types, i.e. $\sigma_k^2 = \sigma^2$. Furthermore, we assume the errors are uncorrelated across cell types. Under these assumptions, the PVE of the eQTL on bulk gene expression is given by:

$$\mathrm{PVE} = \frac{p_c^2 \beta_c^2}{p_c^2 \beta_c^2 + \sigma^2 \sum_k p_k^2}. \tag{7}$$

With out loss of generality, we assume $\sigma^2 = 1$, in other words, our effect sizes are measured at the scale/unit of $\sigma$. Now given $p_k$ of all cell types, and $\beta_c$, we can evaluate PVE. To assess the power, we use this simple relationship from standard linear regression: let $\hat{z}$ be the $Z$-score of the association test, $n$ sample size, then $\hat{z} \sim N(\sqrt{n \cdot \mathrm{PVE}}, 1)$.

In our simulations, we use cell type proportions similar to those in real data. We have 7 cell types with proportions 0.3, 0.2, 0.2, 0.1, 0.1, 0.05 and 0.05, respectively. We consider two cases $p_c = 0.3$ and 0.2, respectively. We then vary $\beta_c$ to assess PVE and then power at $p < 10^{-3}$, using sample size of $n = 500$. To assess how much power is lost from bulk samples, compared with the case where we have "pure" cell type of $c$, we evaluate the power under

$$\mathrm{PVE}_{\max} = \frac{\beta_c^2}{\beta_c^2 + \sigma^2} = \frac{\beta_c^2}{\beta_c^2 + 1}. \tag{8}$$

Our simulation shows that the power of detecting cell-type specific eQTLs, especially when cell type proportion is low, is substantially reduced (Extended Data Fig. 14b).

We note that our assumption of independent gene expression across cell types is probably overly-simplified. In reality, gene expression are likely positively correlated across cell types. Ignoring this correlation most likely leads to under-estimation of the total residual variance of gene expression. For a given eQTL effect size, this under-estimation would lead to over-estimated PVE, and hence power. So the power loss for cell-type specific eQTLs is likely even larger than our simulations showed.

In conclusion, our analysis suggests that the power of detecting cell-type specific eQTLs is likely low in bulk-eQTL studies, and this probably contributes to the observed high level of eQTL sharing across tissues, as well as the observation that the bulk eQTLs explain low heritability of complex traits.
